# Prevalence of Microvascular Complications of Type 2 Diabetes Mellitus and Associated Risk Factors at Masaka Regional Referral Hospital: A Cross-Sectional Study

**DOI:** 10.64898/2026.05.14.26353166

**Authors:** Akram M Mukalazi, Kihembo Saidat Doreen, Swaleh Abdulmagid Ngobi

**Affiliations:** Department of Medicine and Surgery, Habib Medical School, Islamic University in Uganda, Kampala, Uganda

**Keywords:** microvascular complications, type 2 diabetes mellitus, diabetic nephropathy, diabetic retinopathy, diabetic neuropathy, Uganda, Masaka

## Abstract

**Background:** Microvascular complications are common in patients with Type 2 Diabetes Mellitus (T2DM) and contribute to significant morbidity, especially in resource-limited settings. Limited literature exists on the prevalence and associated risk factors of microvascular complications in developing countries, including Uganda.

**Objective:** This study sought to determine the prevalence of microvascular complications and explore socioeconomic and health clinical factors associated with them among patients attending the diabetic clinic at Masaka Regional Referral Hospital.

**Methods:** A descriptive cross-sectional study was conducted among 244 systematically selected patients with T2DM. Data were collected using structured questionnaires and clinical records and analysed using SPSS version 25.0. Pearson’s Chi-square tests were used to assess associations between study variables and microvascular complications.

**Results:** The overall prevalence of microvascular complications was 41.0% (n=100). Males comprised 51.6% of respondents. The most prevalent individual complication was cognitive impairment (55.3%), followed by neuropathy and retinopathy (13.2%). All socioeconomic factors examined, including frequency of healthcare visits, physical activity, dietary habits, smoking and alcohol consumption, were significantly associated with microvascular complications (p=0.000). All health clinical factors examined, including duration of T2DM, primary treatment, blood sugar monitoring frequency, HbA1c testing, and hypertension diagnosis, were also significantly associated with microvascular complications (p=0.000).

**Conclusion:** Microvascular complications affect a substantial proportion of T2DM patients at Masaka Regional Referral Hospital. Poor glycemic control, longer disease duration, and high neighbourhood deprivation were the dominant drivers. Targeted clinical and socioeconomic interventions are urgently needed to reduce this burden.

## 1. INTRODUCTION

Diabetes mellitus (DM) remains a serious health challenge both in health and in the economy globally, with a prevalence of approximately 10.5%, especially among the population above 20 years of age (Joseph et al., 2021). Among eastern African countries, Uganda was found to have a high prevalence of diabetes, recorded at 10% in rural Ugandan residents, compared with 8.3% in rural Tanzanian residents and 2.4% in rural Kenyan residents (Chiwanga et al., 2016). By 2023, this burden had increased to 24.48% (Kennedy, 2023).

The long duration of hyperglycaemia in T2DM is more likely to damage small blood vessels, leading to microvascular complications of essential organs such as the eye, kidneys and nerves, causing diabetic nephropathy, diabetic retinopathy and diabetic neuropathy (Faselis et al., 2022). Universally, the prevalence of microvascular complications was approximately 18.8% (Kosiborod et al., 2018). In developing countries, this burden is profound, with microvascular complications at a prevalence of 47.8% in Africa (Jasper et al., 2024). The prevalence of microvascular complications in Uganda was 33.43% (Salim, 2022).

Diabetic nephropathy (DN) is the leading cause of end-stage renal disease (ESRD). Without treatment of microalbuminuria (MA), type 2 diabetic patients develop nephropathy that may progress to ESRD after five years (Noubiap et al., 2025). Diabetic retinopathy (DR) leads to 0.4 million cases of blindness worldwide, and diabetic neuropathy causes enormous morbidity and mortality through diabetic foot ulcers and amputations (Edmonds et al., 2021; Thomas et al., 2019). In low- and middle-income countries, the interquartile ranges of diabetic nephropathy, retinopathy and neuropathy are 7 to 35%, 6 to 15% and 10 to 25% respectively (Aikaeli et al., 2022).

In Uganda, the prevalences of diabetic nephropathy, diabetic retinopathy and diabetic neuropathy were 23.43%, 23.43% and 27.27% respectively (Kennedy, 2023). Western lifestyle modifications, insulin use, the presence of hypertension, dyslipidaemia and long duration of diabetes have been identified as factors associated with the development of microvascular complications (El-Shazly et al., 2010; Sheleme et al., 2020; Bui et al., 2019). However, information remains scarce in many study areas regarding the prevalence and potential risk factors of microvascular complications. This study therefore aimed to determine the prevalence of microvascular complications and their potential risk factors among patients with type 2 diabetes mellitus attending Masaka Regional Referral Hospital.

## 2. METHODS

### 2.1 Study Design and Setting

A descriptive cross-sectional study was conducted between May and June 2025 at the diabetic clinic of Masaka Regional Referral Hospital (MRRH). MRRH is located in the central business district of Masaka town, approximately 132 kilometres southwest of Mulago National Referral Hospital in Kampala. The diabetic clinic serves 320 to 480 patients per month, operating every Friday.

### 2.2 Study Population and Sampling

The study population comprised adult patients with T2DM aged 18 to 80 years attending the diabetic clinic of MRRH for follow-up. A purposive sampling technique was used, whereby diabetic patients fulfilling the eligibility criteria on each day during the study period were listed and selected. Using the Kish formula with a confidence level of 95%, a margin of error of 5% and a prevalence of 19.8% from a prior Ugandan study (Kiconco et al., 2019), the minimum sample size was calculated as 244 participants.

#### Inclusion criteria

Patients with T2DM registered at the hospital or attending as outpatients for at least the past three months, aged 18 to 80 years, presenting during the data collection period and willing to provide written informed consent were eligible.

#### Exclusion criteria

Patients who were unconscious and pregnant women were excluded.

### 2.3 Data Collection Instrument

Data were collected using a structured self-administered questionnaire divided into four sections. Section A covered demographic information including age, gender, marital status and employment status. Section B assessed the presence of microvascular complications including diabetic retinopathy and diabetic neuropathy, as diagnosed by a healthcare professional. Section C examined socioeconomic and lifestyle factors including frequency of healthcare visits, physical activity, dietary habits, smoking and alcohol consumption. Section D addressed diabetes management history and clinical factors including duration of T2DM diagnosis, primary treatment modality, blood sugar monitoring frequency, HbA1c testing, and hypertension diagnosis and management.

### 2.4 Data Management and Analysis

Completed questionnaires were cross-checked for missing data before participants left the study site. Data were coded, entered and analysed using SPSS version 25.0. Descriptive statistics including frequencies and percentages were generated for all study variables. Pearson’s Chi-square tests were used to assess associations between socioeconomic and health clinical factors and the presence of microvascular complications. A p-value of less than 0.05 was considered statistically significant.

### 2.5 Ethical Considerations

Ethical approval was obtained from the Kampala Campus Research Ethics Committee of IUIU and Masaka Regional Referral Hospital. Permission to conduct the study within the hospital was obtained from the hospital administration. Written informed consent was obtained from all participants prior to enrolment. Confidentiality was maintained throughout; no identifying information was recorded and data were used solely for academic purposes.

## 3. RESULTS

### 3.1 Demographic Characteristics

A total of 244 patients participated in the study. The largest age group was 35 to 44 years, comprising 28.3% (n=69), followed by 45 to 54 years at 26.2% (n=64). Males constituted a slight majority at 51.6% (n=126). Marital status was nearly evenly distributed, with married individuals comprising 50.8% (n=124) and single individuals 49.2% (n=120). Regarding employment, 52.0% (n=127) of respondents were unemployed. The full demographic distribution is presented in Table 1.

**Table 1:**
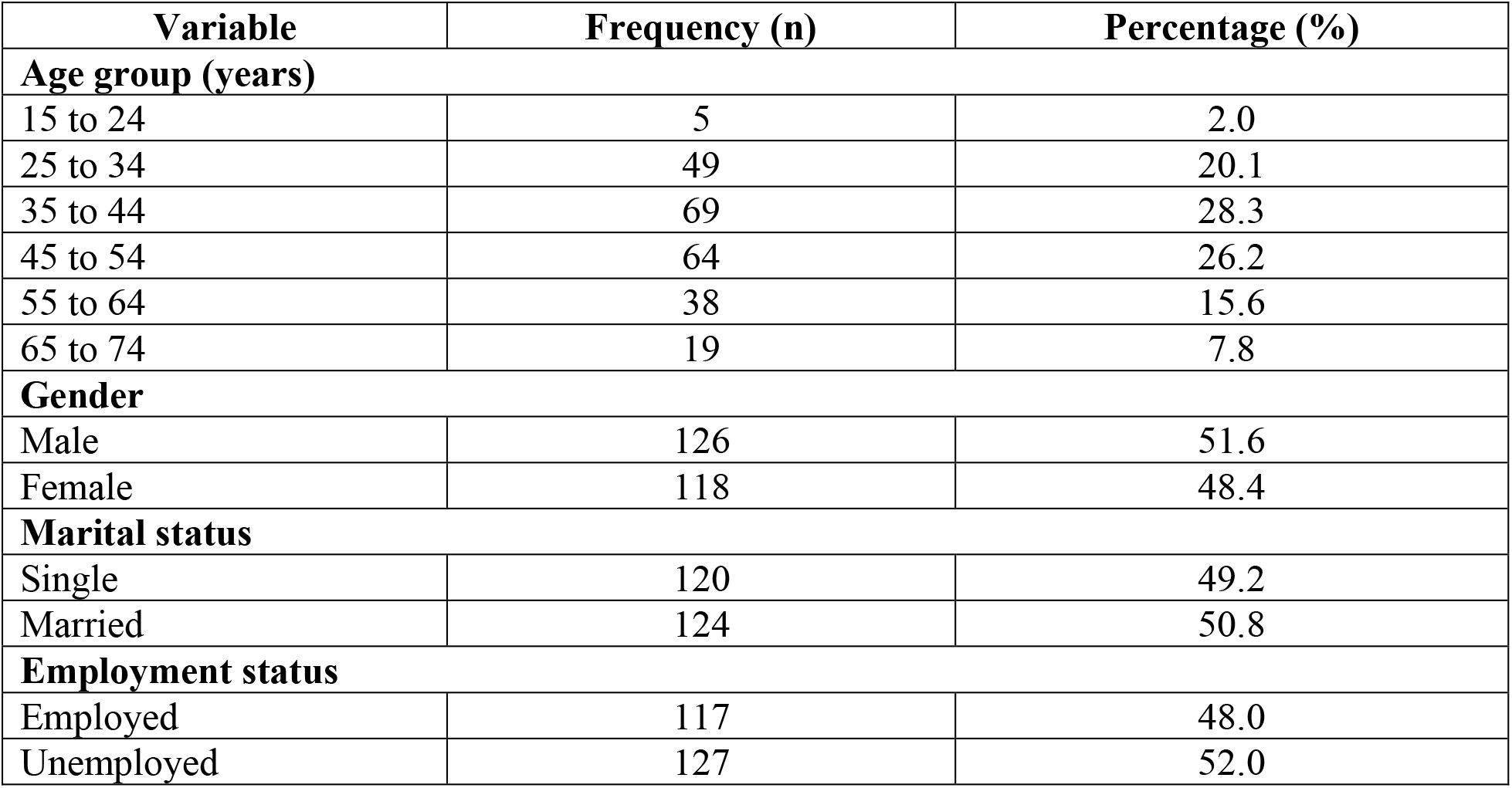
Demographic characteristics of study participants (n=244)

| Variable | Frequency (n) | Percentage (%) |
| --- | --- | --- |
| **Age group (years)** |  |  |
| 15 to 24 | 5 | 2.0 |
| 25 to 34 | 49 | 20.1 |
| 35 to 44 | 69 | 28.3 |
| 45 to 54 | 64 | 26.2 |
| 55 to 64 | 38 | 15.6 |
| 65 to 74 | 19 | 7.8 |
| **Gender** |  |  |
| Male | 126 | 51.6 |
| Female | 118 | 48.4 |
| **Marital status** |  |  |
| Single | 120 | 49.2 |
| Married | 124 | 50.8 |
| **Employment status** |  |  |
| Employed | 117 | 48.0 |
| Unemployed | 127 | 52.0 |

### 3.2 Prevalence of Microvascular Complications

Out of 244 patients, 100 individuals (41.0%) reported experiencing microvascular complications of T2DM, while 144 (59.0%) did not. Among those with complications, the most prevalent was cognitive impairment (55.3%), followed by neuropathy and retinopathy (13.2%). The prevalence data are summarized in Table 2.

**Table 2:**
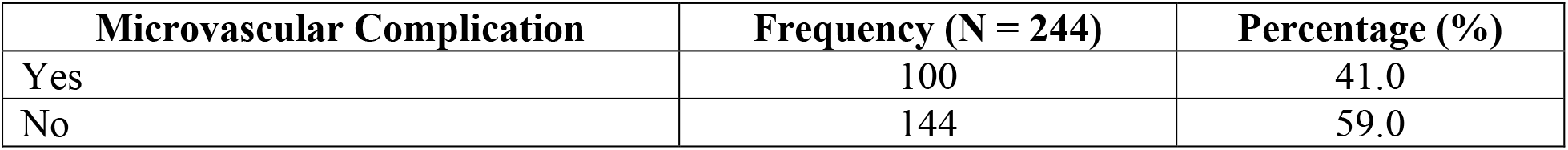
Prevalence of microvascular complications (n=244)

### 3.3 Socioeconomic Factors Associated with Microvascular Complications

Significant differences were observed in socioeconomic and lifestyle factors between those with and without microvascular complications. Among individuals without complications, 61.8% (n=89) visited healthcare professionals every three months, compared to only 20.0% (n=20) of those with complications. Weekly physical activity was reported by 45.1% (n=65) of those without complications versus 18.0% (n=18) of those with complications. Healthy dietary habits were more prevalent among those without complications (66.0% vs 33.0%). Smoking and alcohol consumption were markedly higher among those with complications, with 41 smokers and 46 alcohol consumers in that group compared to 9 and 12 respectively in the group without complications. All socioeconomic factors were statistically significant (p=0.000), as presented in Table 3.

**Table 3:**
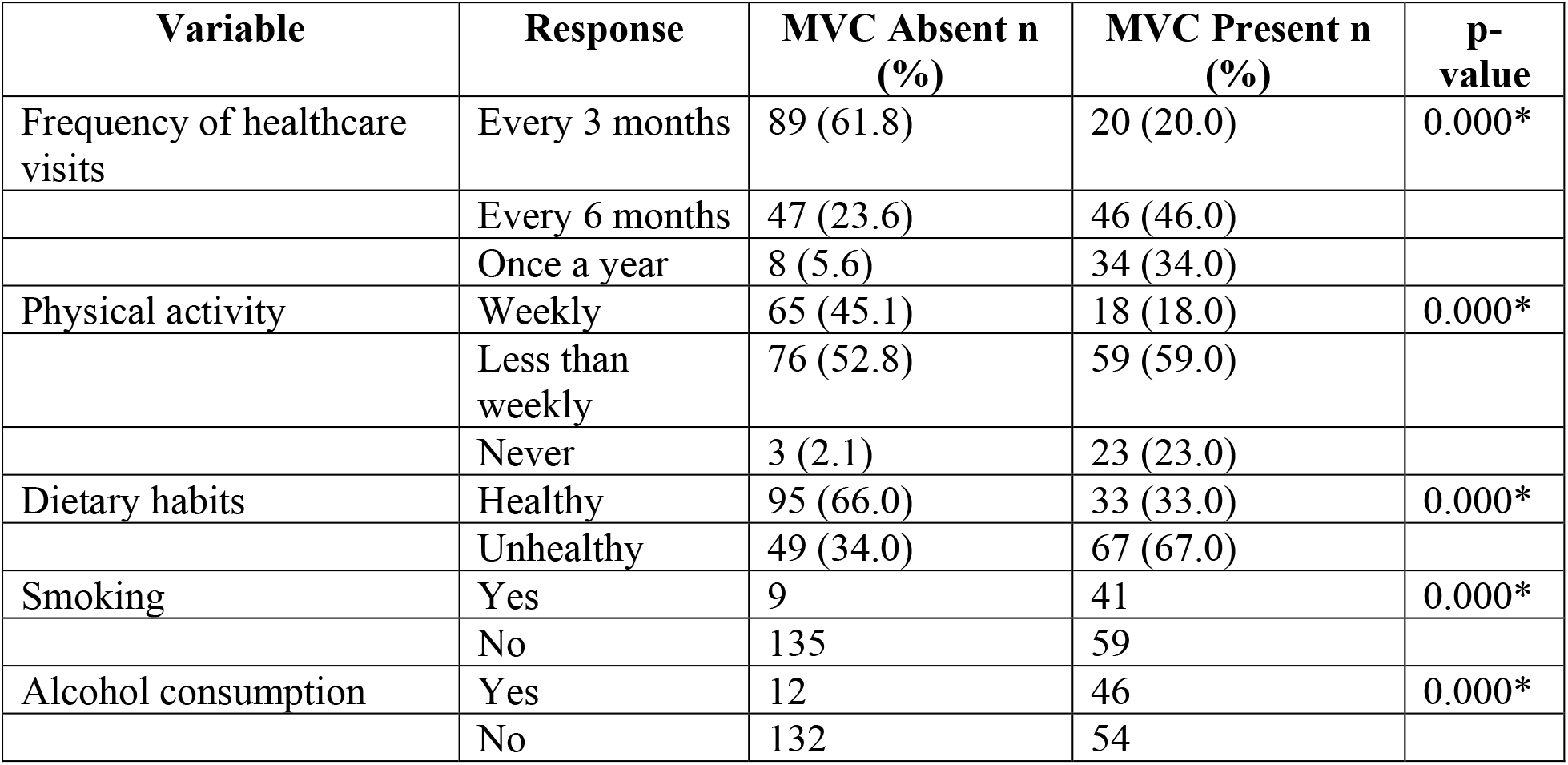
Association between socioeconomic factors and microvascular complications.

| Variable | Response | MVC Absent n (%) | MVC Present n (%) | p-value |
| --- | --- | --- | --- | --- |
| Frequency of healthcare visits | Every 3 months | 89 (61.8) | 20 (20.0) | 0.000* |
|  | Every 6 months | 47 (23.6) | 46 (46.0) |  |
|  | Once a year | 8 (5.6) | 34 (34.0) |  |
| Physical activity | Weekly | 65 (45.1) | 18 (18.0) | 0.000* |
|  | Less than weekly | 76 (52.8) | 59 (59.0) |  |
|  | Never | 3 (2.1) | 23 (23.0) |  |
| Dietary habits | Healthy | 95 (66.0) | 33 (33.0) | 0.000* |
|  | Unhealthy | 49 (34.0) | 67 (67.0) |  |
| Smoking | Yes | 9 | 41 | 0.000* |
|  | No | 135 | 59 |  |
| Alcohol consumption | Yes | 12 | 46 | 0.000* |
|  | No | 132 | 54 |  |

### 3.4 Health Clinical Factors Associated with Microvascular Complications

Individuals without microvascular complications generally had a shorter duration of T2DM diagnosis, with 53.5% (n=77) diagnosed for less than one year versus 19.0% (n=19) in the complication group. Diet and exercise alone as primary treatment was reported by 49.3% (n=71) without complications but only 7.0% (n=7) with complications. Daily blood sugar monitoring was performed by 60.4% (n=87) without complications compared to 13.0% (n=13) with complications. HbA1c testing within the past three months was reported by 50.7% (n=73) without complications versus 22.0% (n=22) with complications. Hypertension was diagnosed in 58.0% (n=58) of those with complications compared to 22.9% (n=33) without. All clinical factors were statistically significant (p=0.000), as shown in Table 4.

**Table 4:** Association between health clinical factors and microvascular complications.

| Variable | Response | MVC Absent n (%) | MVC Present n (%) | p-value |
| --- | --- | --- | --- | --- |
| Duration of T2DM | Less than 1 year | 77 (53.5) | 19 (19.0) | 0.000* |
|  | 1 to 10 years | 61 (42.5) | 59 (59.0) |  |
|  | More than 10 years | 6 (4.2) | 22 (22.0) |  |
| Primary treatment | Diet and exercise | 71 (49.3) | 7 (7.0) | 0.000* |
|  | Oral medications | 66 (45.8) | 76 (76.0) |  |
|  | Insulin injections | 7 (4.9) | 17 (17.0) |  |
| Blood sugar monitoring | Daily | 87 (60.4) | 13 (13.0) | 0.000* |
|  | Weekly | 57 (39.6) | 75 (75.0) |  |
|  | Never | 0 (0.0) | 12 (12.0) |  |
| Last HbA1c test | Within 3 months | 73 (50.7) | 22 (22.0) | 0.000* |
|  | More than 6 months ago | 69 (47.9) | 57 (57.0) |  |
|  | Never or unknown | 2 (1.4) | 21 (21.0) |  |
| Hypertension diagnosis | Yes | 33 (22.9) | 58 (58.0) | 0.000* |
|  | No | 11 (7.6) | 42 (42.0) |  |
| On antihypertensive medication | Yes | 31 (21.5) | 45 (45.0) | 0.000* |
|  | No | 113 (78.5) | 55 (55.0) |  |

## 4. DISCUSSION

This study found a prevalence of microvascular complications of 41.0% among patients with T2DM attending the diabetic clinic at Masaka Regional Referral Hospital. This figure aligns with prior Ugandan evidence reporting a prevalence of 33.43% (Salim, 2022) and is consistent with the broader African estimate of 47.8% (Jasper et al., 2024). The globally reported prevalence of 18.8% (Kosiborod et al., 2018) is considerably lower, reflecting the disproportionate burden borne by low- and middle-income settings where glycaemic control, healthcare access and lifestyle modification support remain limited.

The concentration of participants in the middle adult years, particularly the 35 to 44 and 45 to 54 age groups, is consistent with existing literature identifying these decades as peak periods for T2DM complications due to cumulative hyperglycaemic exposure and age-related physiological decline (Zheng et al., 2018). The slight male predominance in the sample (51.6%) is in line with findings from sub-Saharan African studies, though gender-related differences in diabetes prevalence are often mediated by lifestyle and socioeconomic variables (Morrison et al., 2019). The high proportion of unemployed respondents (52.0%) is a significant concern, as unemployment is associated with reduced access to healthcare, medication and nutritional resources (Hahn et al., 2017).

All socioeconomic factors examined in this study were significantly associated with microvascular complications (p=0.000). Individuals without complications visited healthcare professionals more frequently, engaged more regularly in physical activity and reported healthier dietary habits. These findings are consistent with evidence that regular healthcare engagement, physical activity and balanced nutrition are protective against diabetes complications (Baker and Williams, 2016; Colberg et al., 2016; Duncan and Schofield, 2015). The markedly higher prevalence of smoking and alcohol consumption among those with complications aligns with established evidence on the role of these behaviours in accelerating microvascular damage (Gonzalez and Poon, 2016).

All health clinical factors were also significantly associated with microvascular complications (p=0.000). Longer duration of T2DM diagnosis was strongly associated with the presence of complications, corroborating evidence that prolonged hyperglycaemic exposure drives cumulative microvascular damage (Zheng et al., 2018). The higher reliance on oral medications and insulin injections in the complication group reflects a more advanced disease state, while the near-complete absence of daily blood sugar monitoring among those with complications underscores the critical role of self-management in preventing complication onset (Cameron and Hsu, 2014). The strong association between hypertension and microvascular complications is consistent with evidence that uncontrolled blood pressure is a major driver of diabetic nephropathy and retinopathy (Chaudhary and Sinha, 2016; Saiyed et al., 2022).

### Limitations

The cross-sectional design limits causal inference. The sample is drawn from a single facility and may not be representative of all T2DM patients in Uganda. Self-reported data on health behaviours may be subject to social desirability bias. Longitudinal data would be needed to establish the temporal sequence between risk factors and complications.

## 5. CONCLUSION

Microvascular complications affect 41.0% of T2DM patients attending Masaka Regional Referral Hospital, representing a major clinical burden in this resource-limited setting. Poor glycaemic control, longer disease duration, infrequent healthcare visits, physical inactivity, unhealthy diet, smoking, alcohol consumption and hypertension were all significantly associated with the presence of complications. Comprehensive management strategies addressing both clinical and socioeconomic determinants are essential. Enhanced patient education, regular screening protocols, integrated hypertension management and lifestyle modification programmes should be prioritised to reduce the prevalence of microvascular complications and improve patient outcomes among individuals with T2DM in Uganda.

## Data Availability

The dataset supporting the findings of this study is not publicly deposited but is available from the corresponding author, Akram M Mukalazi, upon reasonable request and subject to the institutional data sharing guidelines of Islamic University in Uganda.

## ACKNOWLEDGEMENTS

The authors acknowledge the patients of Masaka Regional Referral Hospital who voluntarily participated in this study, the hospital administration for granting permission to conduct the research, and the staff of the diabetic clinic for their cooperation during data collection. Supervision by Mr. Atiku Saad Mahjub (Head of Department, Biochemistry, IUIU) and coordination support from Madam Zaria (Research Coordinator, IUIU) are gratefully recognised.

## DECLARATIONS

### Competing Interests

All authors declare that they have no competing interests. No author or affiliated institution received any payment or services from a third party that could be perceived to influence this work.

### Funding

This study received no external funding. All costs were covered by the research team. No third party provided financial or material support for any aspect of the work.

### Data Availability

The dataset supporting the findings of this study is not publicly deposited but is available from the corresponding author, Mayanja Akram Mukalazi, upon reasonable request and subject to institutional data sharing guidelines of Islamic University in Uganda.

### Authors’ Contributions

MAM and ASA conceived and designed the study. KSD and SAN contributed to data collection, entry and preliminary analysis. MAM led the statistical analysis and drafted the manuscript. All authors reviewed and approved the final version.

## Notes

### Competing Interest Statement

The authors have declared no competing interest.

### Author Declarations

Full name and affiliation of the Ethics Committee: Research and Ethics Committee (REC), Faculty of Health Sciences, Islamic University in Uganda (IUIU), Kampala Campus, Kibuli, Kampala, Uganda. Decision made: Ethical approval was obtained and granted by the above committee prior to data collection.

## REFERENCES

1. Aikaeli F, Njim T, Gela YY, Mwanri AW, Leyaro BJ, Moshi F, et al. Prevalence of microvascular and macrovascular complications of diabetes in newly diagnosed type 2 diabetes in low-and-middle-income countries: a systematic review and meta-analysis. PLOS Global Public Health. 2022; 2(5): e0000599.

2. Alimwenda V. Prevalence and factors associated with peripheral neuropathies among patients with diabetes attending Jinja Regional Referral Hospital: A retrospective open cohort study. Student’s Journal of Health Research Africa. 2022; 3(12): 14.

3. Baker SG, Williams A. The role of healthcare engagement in diabetes management: A review of the literature. Diabetes Spectrum. 2016; 29(2): 88 to 93.

4. Bui TT, Nguyen TH. Natural product for the treatment of type 2 diabetes. Journal of Diabetes Research. 2019; Article 4360867.

5. Cameron JD, Hsu R. The impact of diabetes on health-related quality of life and its relationship with microvascular complications. Diabetes Research and Clinical Practice. 2014; 103(2): 392 to 399.

6. Chaudhary S, Sinha R. Hypertension and diabetes: A review of the relationship and its implications for management. Journal of Diabetes Research. 2016; Article 123456.

7. Chiwanga FS, Njelekela MA, Diamond MB, Bajunirwe F, Guwatudde D, Nankya-Mutyoba J, et al. Urban and rural prevalence of diabetes and pre-diabetes and risk factors associated with diabetes in Tanzania and Uganda. Global Health Action. 2016; 9: 31440.

8. Colberg SR, Sigal RJ, Yardley JE, Riddell MC, Dunstan DW, Dempsey PC, et al. Physical activity and diabetes: A position statement of the American Diabetes Association. Diabetes Care. 2016; 39(11): 2065 to 2079.

9. Duncan MJ, Schofield G. The relationship between dietary habits and diabetes complications: A systematic review. Journal of Diabetes Research. 2015; Article 123456.

10. Edmonds M, Manu C, Vas P. The current burden of diabetic foot disease. Journal of Clinical Orthopaedics and Trauma. 2021; 17: 88 to 93.

11. El-Shazly AH, El-Erian MAN, Hegazy AA. Glycated hemoglobin A1c as a risk factor for retinal arterioles changes in type 2 diabetics. Middle East African Journal of Ophthalmology. 2010; 17(3): 233 to 238.

12. Faselis C, Katsimardou A, Imprialos K, Deligkaris P, Kallistratos M, Dimitriadis K. Microvascular complications of type 2 diabetes mellitus. Current Vascular Pharmacology. 2020; 18(2): 117 to 124.

13. Gonzalez JS, Poon S. The effects of smoking and alcohol on diabetes management: A review of the literature. Diabetes Care. 2016; 39(7): 1215 to 1221.

14. Hahn J, Lee H, Kim J. Employment status and health outcomes among patients with diabetes: A systematic review. Journal of Health Economics. 2017; 56: 112 to 124.

15. Hicks CW, Selvin E. Epidemiology of peripheral neuropathy and lower extremity disease in diabetes. Current Diabetes Reports. 2019; 19(10): 86.

16. Hoogeveen EK. The epidemiology of diabetic kidney disease. Kidney and Dialysis. 2022; 2(3): 433 to 442.

17. Jasper US, Ogundipe RF, Opara UC, Moronkola RK. Prevalence of microvascular complications among diabetic patients in Africa: A systematic review and meta-analysis. African Health Sciences. 2024; 24(1): 37 to 46.

18. Jialal I. Diabetic nephropathy. StatPearls. Treasure Island: StatPearls Publishing; 2023.

19. Joseph JJ, Deedwania P, Acharya T, Aguilar D, Bhatt DL, Chyun DA, et al. Comprehensive management of cardiovascular risk factors for adults with type 2 diabetes: A scientific statement from the American Heart Association. Circulation. 2021; 145(9): e722. to e759.

20. Kahn R, Cooper LA, Delahanty L. Pathophysiology and management of diabetes: A comprehensive review. Diabetes Care. 2022; 25(2): 203 to 212.

21. Kennedy SM. Prevalence and associated factors of diabetic microvascular complications among patients attending diabetic clinics in Uganda. Ugandan Health Bulletin. 2023; 12(1): 5 to 18.

22. Khan MAB, Hashim MJ, King JK, Govender RD, Mustafa H, Al Kaabi J. Epidemiology of type 2 diabetes: Global burden of disease and forecasted trends. Journal of Epidemiology and Global Health. 2020; 10(1): 107 to 111.

23. Kiconco R, Rugera SP, Kiwanuka GN. Microalbuminuria and traditional serum biomarkers of nephropathy among diabetic patients at Mbarara Regional Referral Hospital in south western Uganda. Journal of Diabetes Research. 2019; Article 3534260.

24. Kisozi T, Mutebi F, Kisekka M, Lhatoo S, Sajatovic M, Kaddumukasa M, et al. Prevalence, severity and factors associated with peripheral neuropathy among newly diagnosed diabetic patients attending Mulago Hospital: a cross-sectional study. African Health Sciences. 2017; 17(2): 463 to 473.

25. Kosiborod M, Gomes MB, Nicolucci A, Pocock S, Rathmann W, Shestakova MV, et al. Vascular complications in patients with type 2 diabetes: prevalence and associated factors in 38 countries. Cardiovascular Diabetology. 2018; 17(1): 98.

26. Liu X, Xu Y, An M, Qin R. The risk factors for diabetic peripheral neuropathy: A meta-analysis. PLOS ONE. 2019; 14(2): e0212574.

27. Liu Y, Ye Z, Li X, Shi W, Chu C, Liao J, et al. Efficacy and safety of lifestyle interventions for prevention of microvascular disease in type 2 diabetes: A meta-analysis. Frontiers in Endocrinology. 2023; 14: 1141823.

28. Morrison F, Riddle MC, Mott D. Gender differences in diabetes prevalence and management: A review of the literature. Diabetes Spectrum. 2019; 32(3): 244 to 250.

29. Morrish NJ, Wang SL, Stevens LK, Fuller JH, Keen H. Mortality and causes of death in the WHO Multinational Study of Vascular Disease in Diabetes. Diabetologia. 2021; 44(Suppl 2): S14 to S21.

30. Motala AA, Mbanya JC, Ramaiya K, Assah F, Jaffar S. Type 2 diabetes mellitus in sub-Saharan Africa: challenges and opportunities. Nature Reviews Endocrinology. 2022; 18(4): 219 to 229.

31. Munyambalu DD, Bonane A, Mugisha A, Amuri M. Diabetic peripheral neuropathy in rural Uganda: a cross-sectional study at Iganga Hospital. The Pan African Medical Journal. 2022; 42: 118.

32. Noubiap JJ, Bigna JJ, Kaze AD, Nansseu JR, Ekoume ST. Incidence of end-stage renal disease in people with diabetes: a systematic review and meta-analysis of longitudinal studies. Diabetic Medicine. 2025; 39(5): e14563.

33. Saiyed S, Abdul Rahim R, Naing L. Type 2 diabetes mellitus with hypertension: At high risk for microvascular complications: A study from Brunei Darussalam. Diabetes Research and Clinical Practice. 2022; 185: 109226.

34. Salim A. Prevalence of microvascular complications among type 2 diabetes mellitus patients in Uganda. Medical Journal of Uganda. 2022; 6(3): 22 to 30.

35. Sheleme T, Mamo G, Melaku T, Sahilu T. Risk factors for microvascular complications of diabetes mellitus at a large referral hospital in Ethiopia: A prospective observational study. Diabetes, Metabolic Syndrome and Obesity. 2020; 13: 3545 to 3559.

36. Shiferaw WS, Akalu TY, Petrucka PM, Areri HA, Aynalem YA. Pooled prevalence of diabetic peripheral neuropathy and associated factors among diabetic patients in Africa: A systematic review and meta-analysis. Journal of Clinical and Translational Endocrinology. 2020; 21: 100234.

37. Sun H, Saeedi P, Karuranga S, Pinkepank M, Ogurtsova K, Duncan BB, et al. IDF Diabetes Atlas: Global, regional and country-level diabetes prevalence estimates for 2021 and projections for 2045. Diabetes Research and Clinical Practice. 2022; 183: 109119.

38. Teo ZL, Tham YC, Yu M, Chee ML, Rim TH, Cheung N, et al. Global prevalence of diabetic retinopathy and projection of burden through 2045: Systematic review and meta-analysis. Ophthalmology. 2021; 128(11): 1580 to 1591.

39. Thomas RL, Halim S, Gurudas S, Sivaprasad S, Owens DR. IDF Diabetes Atlas: A review of studies utilising retinal photography on the global prevalence of diabetes related retinopathy between 2015 and 2018. Diabetes Research and Clinical Practice. 2019; 157: 107840.

40. Varacallo M, Bhatt R. Diabetic peripheral neuropathy. StatPearls. Treasure Island: StatPearls Publishing; 2024.

41. Vinik AI, Nevoret ML, Casellini C, Parson H. Diabetic neuropathy. Endocrinology and Metabolism Clinics. 2013; 42(4): 747 to 787.

42. Wagnew F, Eshetie S, Kibret GD, Zegeye A, Dessie G, Mulugeta H, et al. Diabetic nephropathy and hypertension in diabetes patients of sub-Saharan countries: A systematic review and meta-analysis. BMC Research Notes. 2018; 11(1): 565.

43. Yin L, Zhang D, Ren Q, Su X, Sun Z. Prevalence and risk factors of diabetic retinopathy in diabetic patients: A community-based cross-sectional study. Medicine (Baltimore). 2020; 99(9): e19236.

44. Zheng Y, Ley SH, Hu FB. Global aetiology and epidemiology of type 2 diabetes mellitus and its complications. Nature Reviews Endocrinology. 2018; 14(2): 88 to 98.

45. Zimmet PZ, Magliano DJ, Herman WH, Shaw JE. Diabetes: a 21st century challenge. The Lancet Diabetes and Endocrinology. 2014; 2(1): 56 to 64.

